# Acceptability and satisfaction of contraceptive vaginal rings in clinical studies: a systematic review and narrative synthesis

**DOI:** 10.1101/2021.06.15.21258958

**Authors:** Thérèse Delvaux, Vicky Jespers, Lenka Benova, Janneke van de Wijgert

## Abstract

**Introduction:** Acceptability of and satisfaction with contraceptive methods are paramount for uptake and continuation. In the current context of multipurpose prevention of pregnancy and sexually transmitted diseases /HIV development, it is critical to have a better understanding of acceptability of and satisfaction with the contraceptive vaginal ring (CVR), including sexual satisfaction. The objective of this study was to review the evidence about CVRs acceptability, and users’ general and sexual satisfaction.

**Methods:** We searched PubMed, CINAHL and Web of Science (until December 31th, 2020) and selected original studies documenting actual use of hormonal CVR and explicitly addressing any of the three outcomes.

**Main Results:** Of a total of 1129 records screened, 46 studies were included. Most studies (n=43, 93%) were prospective, conducted in high-income settings (n=35) and reported on NuvaRing^®^ use (n=31). Overall, 27 (59%) studies included a comparison group, 38 (82%) used exclusively quantitative questionnaires, with qualitative only (n=4, 9%) or mixed methods (n=4, 9%) studies being less common. Ease of CVR insertion/removal/reinsertion was high in all setting and improved with time of use, with qualitative studies supporting these findings. When reported, results on continuation of use were mixed and ring-related events were associated with discontinuation. Among NuvaRing^®^ studies, general satisfaction (being satisfied or very satisfied) was between 80 and 90% and tended to mirror continuation. Sexual satisfaction was less commonly reported and results were mixed. Overall, limited information was provided on actual CVR experiences of women (and men) and cultural norms that may affect sexuality and CVR use.

**Conclusion:** Positive aspects of acceptability of and satisfaction with CVRs were reported but continuation rates and ring-related events deserve further study. More information is needed on actual experiences of women using CVRs, relationship aspects, male partner opinions, and contextual norms to better understand the acceptability of and satisfaction with CVRs.

**Key strengths and limitations of this study:**

- This review brings an historical and international perspective on acceptability and satisfaction of contraceptive vaginal ring (CVR), since the 1970’s in high, middle and low income countries.
- An holistic approach was used, including original studies documenting actual use of hormonal CVR and explicitly addressing acceptability, general *and* sexual satisfaction.
- Our results may inform the development and promotion approaches for CVR and more broadly vaginal rings that could provide combined prevention of HIV, other sexually transmitted infections and pregnancy.
- Given the lack of standardized definitions of acceptability and satisfaction, articles documenting CVR acceptability or satisfaction that were not explicitly using this terminology and instead referred to continuation or adherence may have been missed.
- From the methods sections of included papers we could not always deduct whether interviews included open-ended questions. This may have led to under-recording of the use of semi-structured interviews.

## INTRODUCTION

Contraceptive vaginal rings (CVRs) have been developed since 1970 and three CVRs are currently available: the etonogestrel and ethinyl estradiol ring (marketed as NuvaRing^®^), the progesterone ring for breastfeeding women (Progering^®^), and the recently approved segesterone acetate (previously called nestorone) and ethinyl estradiol ring (Annovera^™^ (1,2). Advantages of CVRs are multiple: they are user-initiated and controlled, independent of sexual acts, and can provide long-term effective protection (1). Moreover, vaginal rings could be designed to include several active ingredients that provide prevention for HIV, other sexually transmitted infections, and pregnancy (3).

Acceptability of and satisfaction with contraceptive methods impact uptake, adherence, and continuation, and therefore contribute significantly to contraceptive effectiveness (4). In clinical studies, acceptability of contraceptive methods is often documented through the effect of the product on bleeding patterns/cycle control, its side effects, and the duration of use. Satisfaction tends to reflect the user’s perceptions of the product, and is assessed quantitatively through levels of satisfaction during actual use and/or indirectly assessed through willingness to use in the future or recommend the method (1). Both the concept of acceptability and satisfaction are in fact intertwined as illustrated by validated quantitative tools in which overall satisfaction is considered a dimension of acceptability (5) (6). Moreover, given vaginal administration, CVRs may affect sexual relationships. To this end sexual satisfaction with CVR has been studied more specifically using sexual function assessment tools such as Female Sexual Function Index 19 (FSF 19). In reality, acceptability and satisfaction are complex concepts that are influenced by physical, behavioral, physiological, interpersonal and structural factors. Recent studies documenting the effectiveness of vaginal products and devices in the field of HIV prevention have confirmed the key contribution of acceptability to adherence and theoretical frameworks presenting pathways from various acceptability dimensions towards satisfaction and then to adherence have been developed to aid further inquiry (7).

Given the current focus and importance of multipurpose technology for prevention of pregnancy and STIs/HIV, it is critical to have a better understanding of what is commonly considered as acceptability and satisfaction of CVR, and main reported results regarding these outcomes, including sexual satisfaction. The objectives of this study were to review the overall evidence of contraceptive vaginal rings (CVRs) acceptability, general and sexual satisfaction.

## MATERIAL AND METHODS

### Protocol and registration

The study protocol was registered on PROSPERO (CRD42017079157).

### Literature search

Databases searched were PubMed, CINAHL and Web of Science with a cut-off date of 31 December 2020. The main search terms were “contraceptive vaginal ring” AND “acceptability” OR “satisfaction” OR “sexual satisfaction”, and synonyms of each of these terms were also included. Additional search terms included “qualitative methods“, “mixed-methods” and “trials”. The search strategies were adjusted according to the specifications of each database. Additional relevant publications from other sources (reference lists) were also included (Supplementary Material S1_Search strategy). The PRISMA (Preferred Reporting Items for Systematic Reviews and Meta-Analyses) framework Guidelines, Flow Diagram and checklist were utilized to undertake the review.

### Selection criteria

We considered any type of study design with the exception of reviews and opinion papers. Studies were eligible if they included actual CVR use by healthy women of reproductive age (15-49 years) and *explicitly* addressed acceptability, satisfaction, and/or sexual satisfaction. We did not use specific definitions of acceptability and satisfaction because we wanted to learn which definitions or concepts the various authors had used. Similarly, we did not select studies based on research methods used but excluded reviews and opinion papers or commentaries, validation studies, studies that evaluated non-contraceptive vaginal ring use (such as rings for hormonal replacement therapy), or assessed acceptability or willingness to use hypothetically in the absence of actual user experiences. Studies that only enrolled women with a specific health condition (such as diabetes), and full texts in languages other than English, French, Dutch, Spanish, or Italian were also excluded (n=3). In case of multiple articles presenting data from the same study with the same outcomes of interest, only the primary paper was included in the review.

### Study selection

Each title and abstract was screened by two independent reviewers (TD, VJ) using the inclusion criteria described above. Full texts of all papers selected in title and abstract screening were checked by both reviewers before inclusion and any discrepancies were discussed until consensus was reached.

### Study quality assessment and data synthesis

A standardized pre-tested form was used by TD to extract data from full texts on study characteristics: author names, year of publication, journal, study setting, study design, ring use and comparison group(s) (if any), research methods used and main findings related to acceptability, overall satisfaction, and sexual satisfaction, sample size, randomization process, and presence of a control group related to methodological quality assessment were also extracted but were not considered a core component of this project as we wanted to provide an overview of methods used to document acceptability and satisfaction.

### Patient and Public Involvement

No patient or public involvement took place in the design or conduct of this systematic review which included 46 papers from many countries worldwide.

## RESULTS

Of 1308 publications that were identified through database searching, after removal of duplicates, 1129 titles/abstracts and 96 full texts were reviewed, and 46 articles (primary studies) were included (Figure 1).

**Figure 1.**
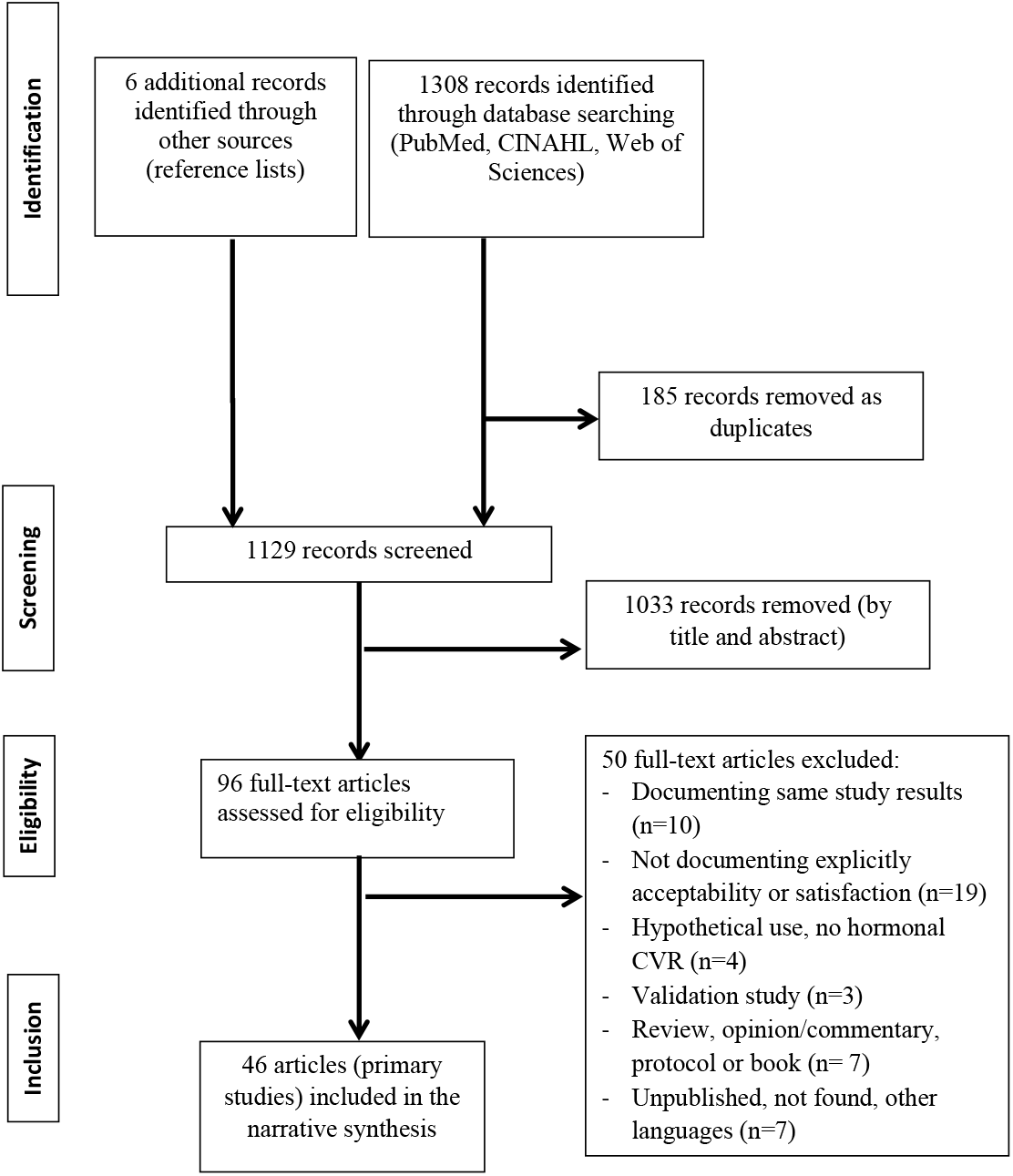
PRISMA Flow chart of the studies selection process for the review on contraceptive vaginal rings, acceptability, general and sexual satisfaction.

### Studies design, methods, characteristics and settings

Nineteen studies (41%) were randomized clinical trials (8–26), 24 were prospective non-randomized studies (27–50), and three were cross-sectional studies (51–53) (Table 1). Twenty seven studies (59%) used a controlled design, comparing CVR users to users of other hormonal methods (such as a combined oral contraceptive pill [COC] or patch) or to non-hormonal contraceptive methods (such as the copper intrauterine device), or comparing users of CVRs containing different hormonal dosages or the same CVR for different durations. The remaining 19 studies did not include a comparison group (Table 1).

**Table 1.** Types of study design, methods, participants and study duration.

| Type of study<br>N (%) | Comparison<br>group<br>N (% per<br>study type) | Quantitative<br>methods<br>Structured<br>Questionnaire | Qualitative<br>methods *<br>Semi-structured,<br>IDI, FGD | Study<br>participants<br>N Range | Study<br>duration<br>N months<br>Range |
| --- | --- | --- | --- | --- | --- |
| Prospective<br>randomized<sup>8-26</sup> **19 (41)** | 19 (100) | 19 (100) | 2 (11) | 14-983 | 1-13<br>months |
| Prospective non<br>randomized<sup>27-50</sup> **24 (53)** | 7 (30) | 21 (88) | 5 (21) | 27-5823 | 2-24<br>months |
| Cross sectional<sup>51-53</sup> **3 (7)** | 1 (33) | 2 (67) | 1 (33) | 32-26,250 | - |
| **Total** **46 (100)** | **27 (59)** | **42 (91)** | **8 (17)** | **14-26,250** | - |

\* Four studies used both quantitative and qualitative methods (mixed-methods); *IDI: In Depth Interview*; *FGD: Focus Group Discussion*

Most studies (42/46, 91%) used quantitative structured questionnaires, and eight studies (17%) used qualitative semi-structured or in depth interviews (IDIs), and/or focus group discussions (FGDs) (24,25,29,35,40,43,48,51) (Table 1). Overall 38 (82%) studies used exclusively quantitative structured questionnaires, while four (9%) studies used only qualitative methods and four studies (9%) used both quantitative and qualitative methods (24,25,35,48). The number of participants in the prospective studies ranged from 14 to 5,823, with on average 50 to 200 participants. One cross-sectional study was larger with up to 26,250 participants. The duration of use in the prospective studies ranged from one to 24 menstrual cycles, with about a third (n=13) covering 12 cycles or more, another third (n=13) covering six to 12 cycles, and the remaining studies covering three cycles or fewer (Table 1).

Studies were performed in one or more high income settings i.e. countries of Europe, USA, Canada, or Australia (n=35/46 studies, 76%), Latin America (n=5), Israel (n=2), Asia (n=5, of which three were in India), and Africa (n=5, including one study each in Rwanda, Kenya, South Africa, and two studies in several (mostly sub-Saharan) African countries) (Table 2).

**Table 2.** Study settings and types of contraceptive vaginal rings.

| Types of contraceptive vaginal ring | *Study settings* |  |  |  |  | All studies<br>N (%) |
| --- | --- | --- | --- | --- | --- | --- |
| | *Europe, USA, Canada, Australia \$* | *Latin America* | *Israel* | *Asia §* | *Africa* | |
|  | *N* | *N* | *N* | *N* | *N* |  |
| **NuvaRing® \$\$** | 24 | - | 2 | 3 | 3 | **31\* (67)** |
| **Annovera™** | 2 | 2 |  |  | 1 | **2\* (4)** |
| **Progering®** | - | 1 |  | - | - | **2 (4)** |
| **Levonorgestrel** | 3 |  |  | 2 | 1 | **4\* (9)** |
| **Levonorgestrel/Estradiol** | 1 | 2 |  |  | - | **3 (7)** |
| **Other CVR** | 4 | - |  |  | - | **4 (9)** |
| **Total, N (%)** | **35 (76)** | **5 (11)** | **2 (4)** | **5 (11)** | **5 (9)** | **46 \* (100)** |

*High income settings: Europe, USA, Canada, Australia and Israel*
*\* "All studies" column values are less than the sum of all column values as some studies include several settings; \$ Any of these settings or several of them; § Including 3 studies in India; \$\$: including one study with a new etonogestrel/ ethinyl estradiol ring (Kirkos®);*

NuvaRing^®^ was the most studied CVR (31/46 studies, including one recent study testing a new etonogestrel/ ethinyl estradiol - Kirkos^**®**^ - against NuvaRing^®^), while two studies evaluated use of the Annovera^™^ ring and two others the Progering^®^ among breastfeeding women. The remaining 11 studies investigated CVRs containing Levonorgestrel (LNG) alone, combined with ethynyl estradiol or other progesterone regimens that were not further developed and did not make it to the market (Table 2).

### Main findings on acceptability, satisfaction and sexual satisfaction

Overall definitions of acceptability and satisfaction or how these outcomes were described, varied across studies, over time and according to the type of CVR. We will therefore present main results on these outcomes by type of CVR.

#### NuvaRing^®^

Studies documenting acceptability and / or satisfaction of NuvaRing^®^ commonly used structured questionnaires assessing the following similar dimensions: clarity of instructions; ease of use (including to insert/remove the ring); ease of package use; compliance or adherence (including removals and spontaneous expulsions); cycle related characteristics (menstrual changes or pain); sexual comfort (whether the ring was felt by the woman or the male partner or whether the partner objected to the ring, without investigating sexual frequency, pleasure or satisfaction) and overall satisfaction. These seven dimensions were included in a validated 21-item questionnaire by Novak et al.(54) and subsequently used in other studies (20,30– 32,36,37,47). The ORTHO-BC-SAT satisfaction questionnaire related to the use of hormonal contraception in general and including eight dimensions similar to Novack et al. questionnaire was used in one CVR study (18). Over 80% of NuvaRing^®^ users in all studies using the seven/eight dimensions showed that CVR instructions and packaging were clear, and that the ring was easy to insert and remove (Table 3). In Kenya, a qualitative study showed that unease with vaginal insertion as well as ring placement issues (slippage, expulsion) created initial challenges requiring clinician assistance and practice for some participants (29). Similarly, an in-depth discussion with users in Rwanda showed that initial worries regarding CVR insertion reduced over time with actual ring use and ring insertions and removals were henceforth described as easy (25). On the other hand, two studies reported that the previous use of tampons did not seem to influence satisfaction or successful ring use of CVR(13,51). A number of studies reported spontaneous expulsions rates ranging from less than 2% (14,16) to 5-20% (20,35,45,50). A study in Switzerland showed that 17.5% of adverse events were ring related, such as feeling the ring, vaginal discomfort, vaginal expulsions (30). In a study in The Netherlands (37), women who felt the ring were more likely to remove it (sometimes, regularly, always) during intercourse compared to those who did not feel it (22% vs. 6%), Table 3. An in-depth study among adolescents in the US using NuvaRing^®^ revealed that five of 32 participants discontinued because of ring-related events (51). In Kenya, minor side-effects were described, and concerns centered on ring efficacy, negative effect on a woman’s sexual desire, and future fertility issues, and non-suppression of menstruation which was favored by most participants (29). In Rwanda, most women reported and appreciated the absence of negative side effects (25).

**Table 3.**
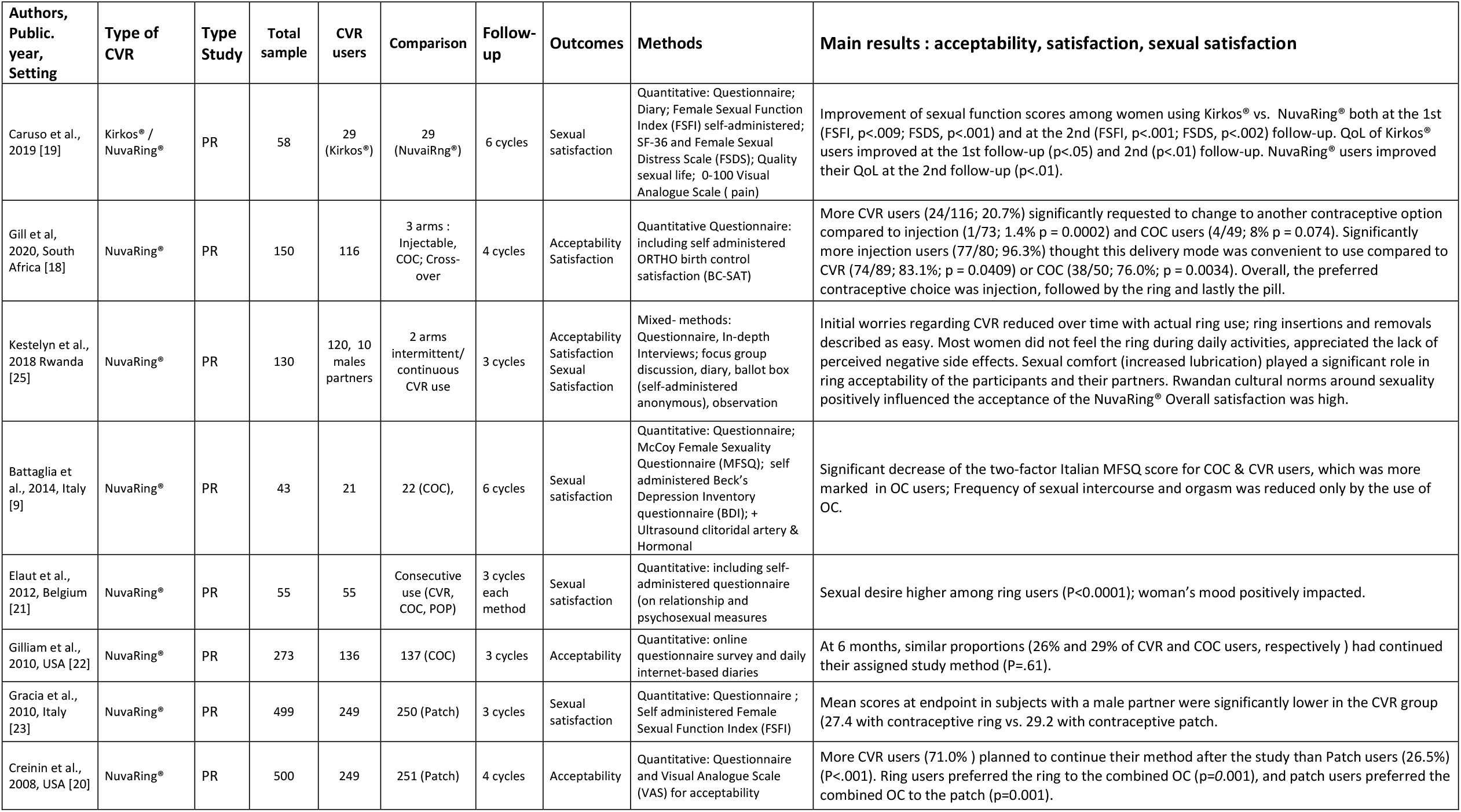
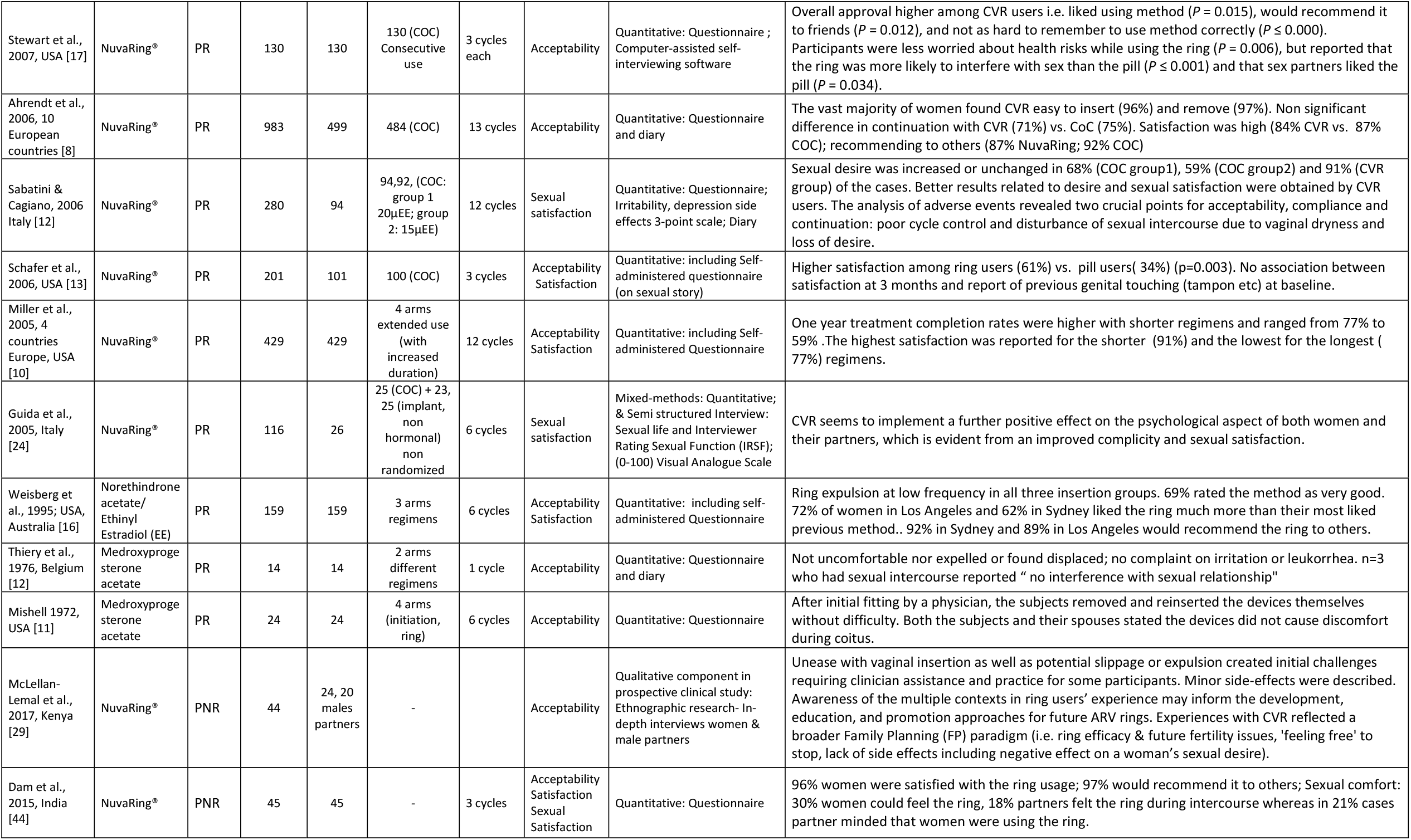
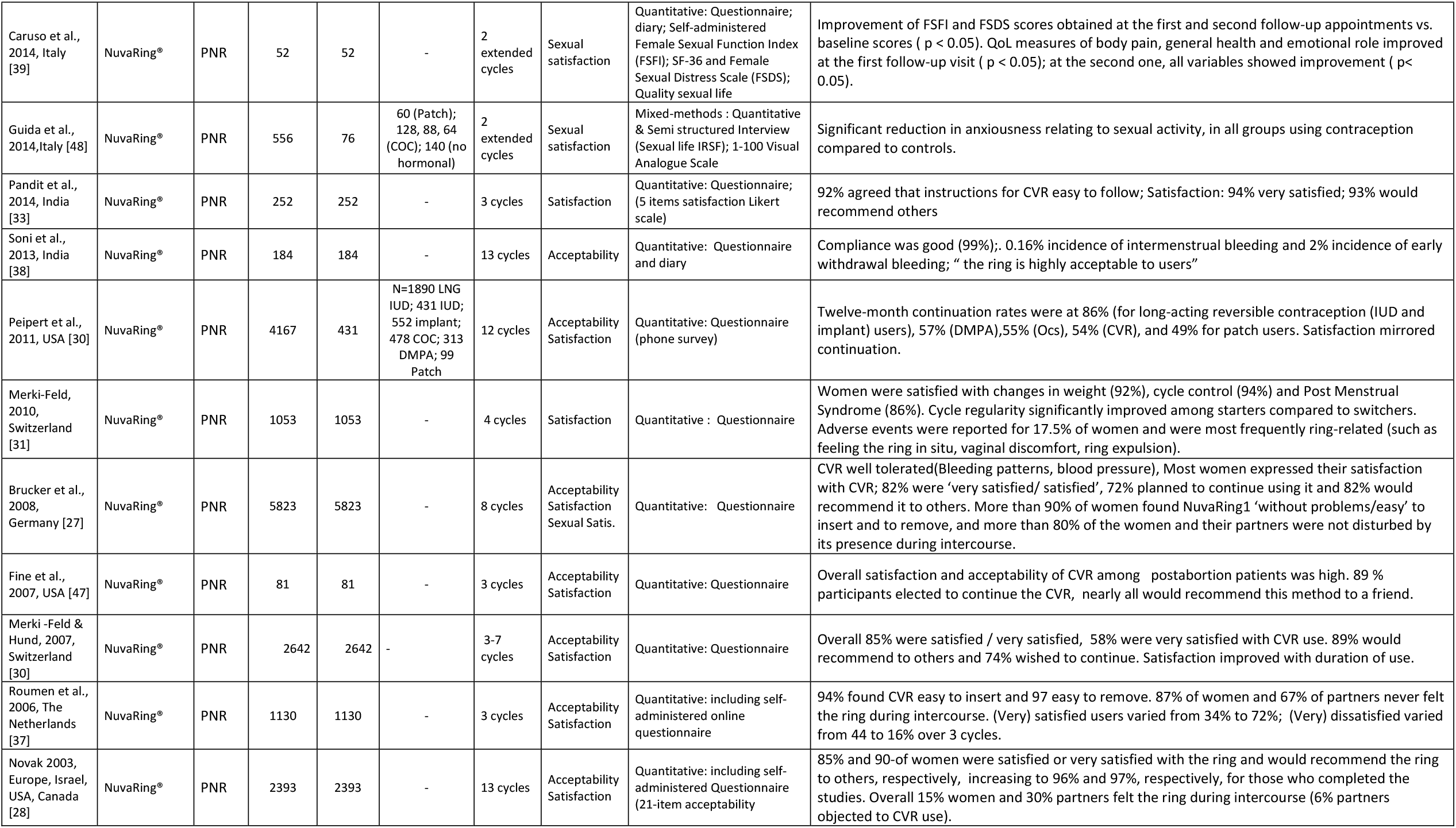
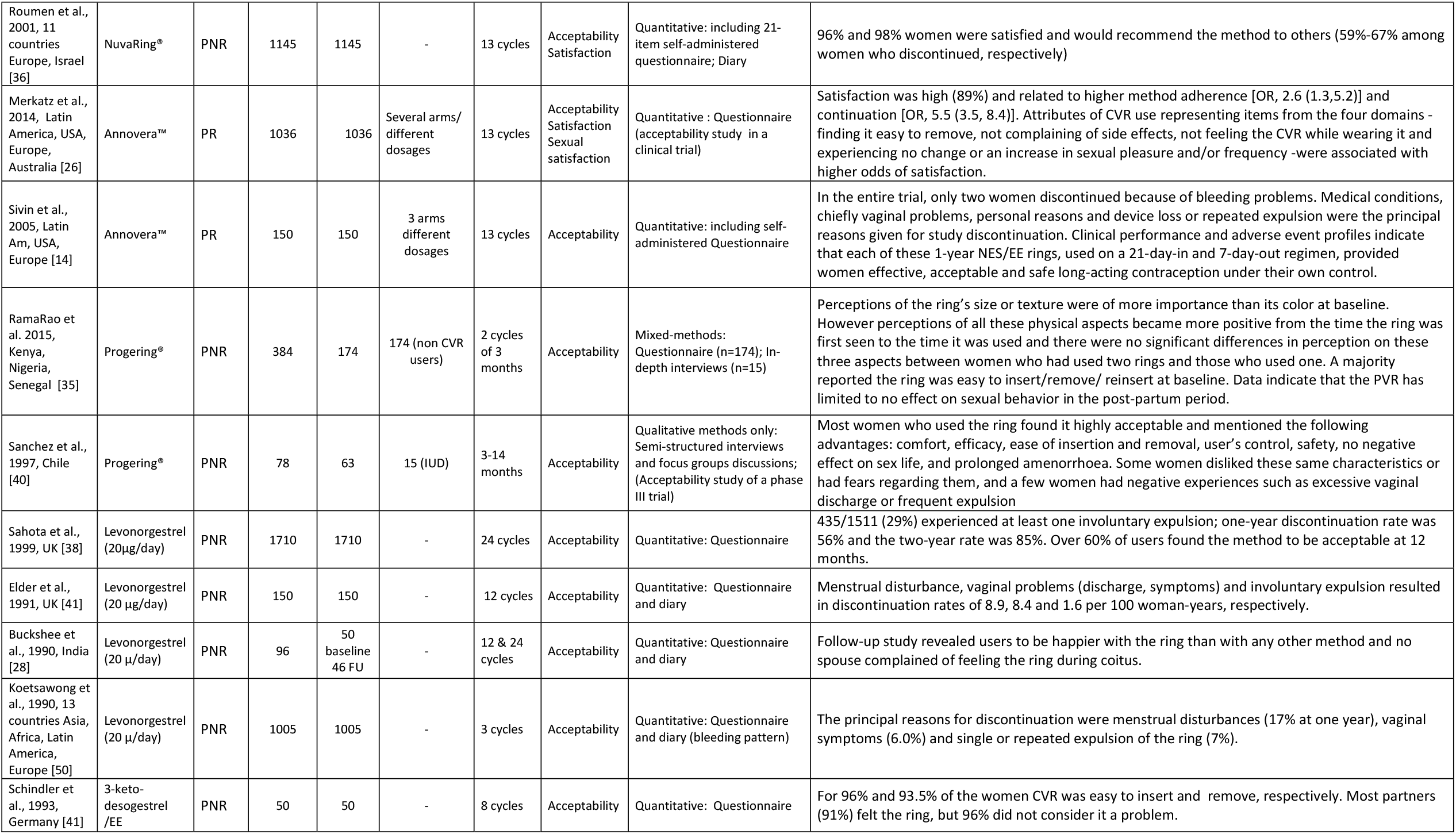
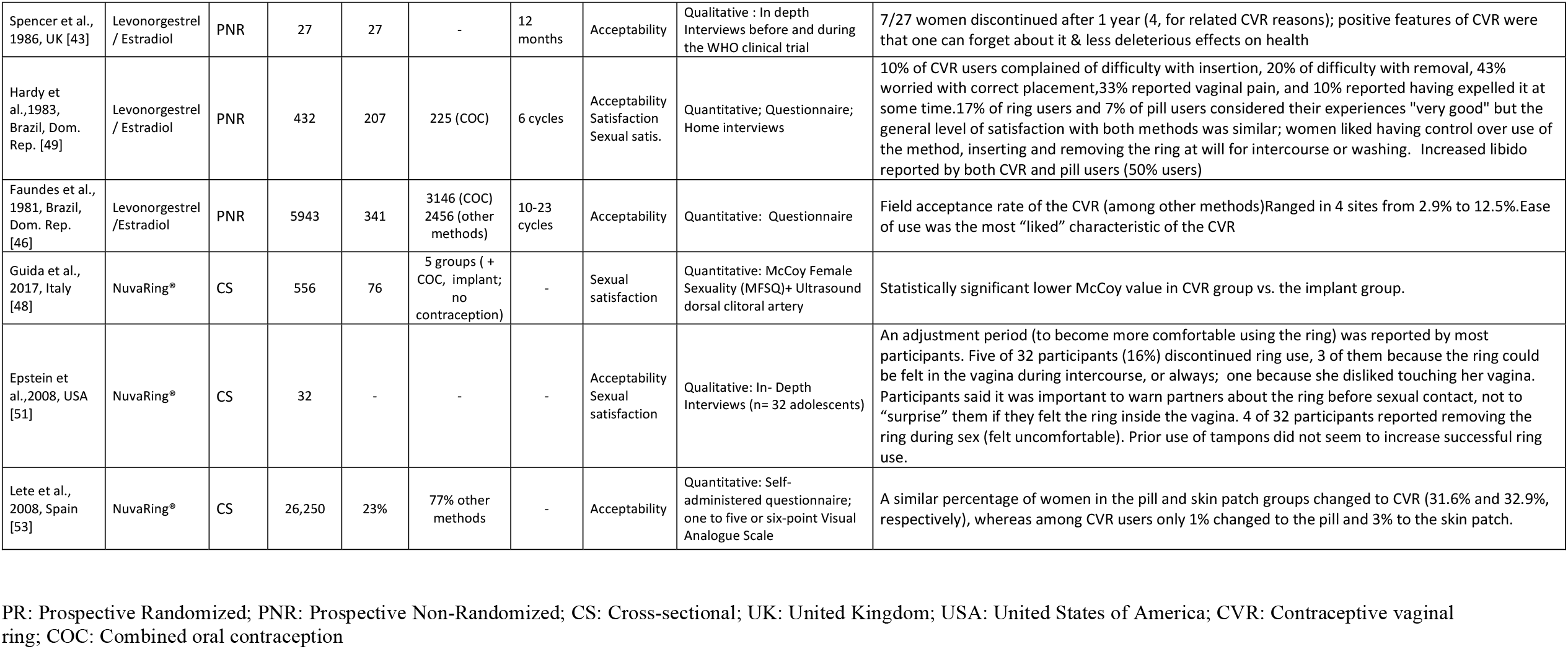
Study characteristics, type of CVR and study, participants, comparison group, outcome(s), methods used, and main results, presented by type of CVR then chronologically.

Results regarding continuation rates reported in acceptability/ satisfaction studies were mixed. Some NuvaRing^®^ studies showed a higher willingness to continue the use of the method (71% for CVR versus 26.5 for Patch users (20) or a higher continuation rate (1% CVR changed to pill or patch vs. 32-33% COC and Patch who changed to CVR (20,53). One year treatment completion rates were higher (77%) with shorter NuvaRing^®^ treatment regimens compared to one-year extended regimen use (59%)(10). However, other studies showed similar (high or low) continuation rates compared to COC (71% CVR vs. 75% COC (8,22); 26% CVR vs. 29% COC) (9). Twelve-month continuation rates were at 54% for CVR users compared to 86% among long-acting reversible contraception (IUD and implant), 57% for DMPA, 55% for COC, and 49% for patch users in a study conducted in the US. A recent study conducted among adolescents in South Africa showed that more NuvaRing^®^ users (24/116; 20.7%) significantly requested to change to another contraceptive option compared to injection (1/73; 1.4% p = 0.0002) and COC users (4/49; 8% p = 0.074) (18).

Overall satisfaction with NuvaRing^®^, measured on a 4-6 point Likert scale or using a dichotomous variable (Yes/No), ranged between 80% and 90%, with studies without a comparison group (other contraceptive method) more tending to report the highest satisfaction rates (Table 3). However, a high level of satisfaction (“being very satisfied”) varied across studies and ranged between 30% and 94% (the highest proportion reported in a study conducted in India) (13,31,33,37). In Rwanda, general satisfaction with NuvaRing^®^ was high (over 80%) and concurred with qualitative findings and a ballot box (anonymous) survey at the end of the trial (25). In South Africa, significantly more injection users (96.3%) thought this delivery mode was convenient to use compared to NuvaRing^®^ (83.1%; p = 0.0409) or COC (76.0%; p = 0.0034). Overall, the preferred contraceptive choice was injection, followed by the ring and lastly the pill (18). Willingness to recommend NuvaRing^®^ to others ranged from 60% to over 90% in the studies documenting satisfaction.

A number of studies reported ‘sexual comfort’ as whether NuvaRing^®^ was felt by partners (up to 30%), whether a number of partners found it bothersome or did mind (5-20%) (27,30,36,44), objected its use (6%) (32) or did prefer the pill (5-30%) instead of CVR (17). A multicenter NuvaRing^®^ study in high income settings pointed out that sexual comfort for the women who prematurely discontinued participation in the studies was only marginally lower than for those who completed them (32).

Sexual satisfaction while using NuvaRing^**®**^ was reported in a total in 16 studies, exclusively (n=9) or with acceptability and general satisfaction (n=7). Through the use of female sexual function indexes, scales or diaries, these studies showed mixed sexual satisfaction results with NuvaRing^®^ use. Two studies (one without comparison group, another one comparing a new CVR Kirkos^®^ to NuvaRing^®^) conducted in Italy reported an improvement of all variables between baseline and follow up (39). Two prospective controlled studies conducted by Guida et al. also in Italy reported improved overall and sexual relationship (“complicity”) among couples (26) and reduced anxiousness compared to COC users (56). Increased sexual desire (compared to COC or progestin only pill was reported among NuvaRing^**®**^ users in a small study in Belgium (21), and increased or unchanged sexual desire in another study comparing NuvaRing to low oestrogen dosed COC (12). In a randomized study in Rwanda, controlling for intermittent vs. continuous NuvaRing^**®**^ use (but not for another contraceptive method), most women reported that ring use stimulated conversations with their partners about increased lubrication and sexual desire, but also about family planning and more general relationship topics. Most women (81%) reported at least once during ring use that the ring made sex feel better and this increased to 87% at the last study visit. Qualitative data confirmed this finding “This ring should be promoted as a sex enhancer” (25). The authors highlighted that Rwandan cultural norms around sexuality positively influenced the acceptance of the NuvaRing^**®**^. On the other hand, a randomized trial found that the McCoy female sexuality score decreased significantly over treatment among COC and CVR users (9). A recent cross-sectional study reported significantly lower median values of female sexuality indexes in the CVR group compared to implant (52). Two studies found significantly decreased libido (3.3% vs. 0.8%) or mean female sexual function indexes with the ring compared to COC or patch users, respectively (21,31).

Finally, male partner’s opinions about NuvaRing^**®**^ were usually indirectly assessed by asking women about their partners’ perception of the CVR (dimension, sexual comfort). Only two studies interviewed (qualitatively) the male partners themselves on perceptions and experiences with the ring (25,29): In Kenya experiences with CVR reflected a broader family planning (FP) paradigm: FP intentions and disclosure practices were influenced by partner support, socioeconomic factors, religion, cultural beliefs, and societal norms, including female sexuality (29). In Rwanda, finding from a limited number of male partners’ interviews were in line with high acceptability and satisfaction reported by women (25).

#### Annovera™ and Progering^®^

Acceptability of Annovera™ was reported as high in two studies. Similar acceptability dimensions than in NuvaRing^**®**^ studies were used in an Annovera™ trial in Europe, USA and Latin America and included in a theoretical framework presenting a pathway from acceptability to satisfaction then further to adherence and continuation (26). In the same study, satisfaction with Annovera™ was rated high (89%) and was associated to adherence and continuation (p<0,001). Not feeling the ring while wearing it and experiencing no change or an increase in sexual pleasure and/or frequency was associated with higher odds of satisfaction (Table 3) (26). An earlier study showed an overall one-year continuation rates at 73%. Medical conditions, mainly vaginal problems, personal reasons and device loss or repeated expulsion were the principal reasons given for study discontinuation. (14).

Acceptability of the progesterone vaginal ring was rated high including ease to insert/ remove/reinsert in African and Latin American settings and perceptions positively improved between the time the ring was first seen and the time it was used (35,40) (Table 3). Perceptions of the ring’s size or texture were reported of more importance than its color at baseline in African settings (35). In Latin America, 5-30 % of women reported negative experiences (vaginal symptoms-excessive discharge or expulsion) (40) while in African settings expulsion reported rate was 5%. The study in sub-Saharan Africa included “family support” as an additional dimension of acceptability (35) and reported using a theoretical framework including other stakeholders such as health care providers, programme managers and policy makers although the framework was not presented. In the same study, data indicated that the PVR had limited to no effect on sexual behavior in the post-partum period (35).

#### Other types of CVRs

Earlier studies on other types of CVRs reported on acceptability often referring to clinical features and tolerability. Vaginal symptoms, expulsions and menstrual disturbances led to discontinuation among Levonorgestrel 20µg ring users (45,50). Similarly results from a qualitative study with the same ring conducted in the UK (parallelly to a WHO randomized trial) showed that overall seven of 27 women discontinued after a year and four of them for ring related reasons (43). In a study conducted in the early 80’s in Latin America with a Levonorgestrel/ Estradiol ring, 43% of women reported being worried about correct ring placement/ insertion (49) (Table 3).

## DISCUSSION

Many studies using mostly quantitative structured questionnaires have documented acceptability and satisfaction of hormonal CVR, particularly NuvaRing^*®*^. Overall, CVR studies show that easiness to insert/remove/reinsert CVRs was high. Continuation rates, when reported, showed mixed results. Among NuvaRing^®^ studies, general satisfaction (being satisfied or very satisfied) was between 80 and 90% although limited information was provided on actual women’s experiences while using CVR, relationship attributes (such as couple communication and decision to use CVR) and contextual elements such as community perceptions of contraception and the CVR, gender/sexual norms and experience.

Ease of insertion/removal/reinsertion of CVRs was reported in most included studies and rated high, including in Latin American or African settings. Among included studies qualitative data on actual experiences of women while using the ring showed that initial worries related to CVR itself or its use, such as aspect, insertion, removal, feeling the ring inside the vagina improved over time (25,29,51), as reported in one qualitative systematic review of CVR and one systematic review of vaginal rings (55,56). Initial concerns sometimes required additional support from the provider or practice from the user (29,55), or benefited of an adjustment period as reported among adolescents and younger users in the US (51).

User’s perception of the ring were sparsely documented and data on ring expulsions was limited in the identified studies. However, when documented, ring-related reasons (slippage, expulsion, vaginal problems or discomfort) contributed for a proportion of women to discontinuation of all types of CVRs and confirmed findings from previous reviews (55,57). Expulsions and mechanical properties of the ring were included as a specific dimension in acceptability theoretical frameworks that were used in two included studies (25,26) and in vaginal ring HIV prevention studies (7) and deserve to be further addressed in future studies.

When reported, willingness to continue CVR use or continuation rates showed mixed results compared to contraceptive patch or pill users. Some evidence suggests that long acting contraceptive (implants or IUD) have higher continuation rates compared to short acting methods such as COC, but also patch and CVR. In South Africa, injectables showed the highest continuation rates and satisfaction.

Among NuvaRing^®^ studies, general satisfaction (being satisfied or very satisfied) was reported between 80 and 90%. However, as highlighted in our results, a comparison group (including the use of another contraceptive method) was present in about half studies. Data triangulation between quantitative and qualitative data contributed to confirm or provide more information on satisfaction and factors, such as increased lubrification, leading to satisfaction and adherence (25,35,55). Standard clinical trials in the field of CVRs mostly used structured questionnaires to assess acceptability, satisfaction, and/or sexual satisfaction. Mixed-methods approaches combining quantitative and qualitative data collection were less commonly encountered. Clinical trial teams may be less familiar or reluctant to use qualitative approaches, because this requires additional resources, time, and expertise over-and-above those required to carry out a clinical trial. Furthermore, qualitative study designs often use a purposive sampling strategy enrolling small numbers of participants, which is different from clinical trial designs based on representative sampling and statistical power calculations. However, recent clinical trials of vaginal microbicides for HIV prevention have highlighted the importance of incorporating qualitative research into clinical trial designs (58).

‘Sexual comfort’ usually referred to whether the ring was (reported) as felt either by women or partners during sexual intercourse, or if the male partner ‘minded’ the ring or its physical effects during intercourse (33). This issue raised concerns among less than a third of women in all context studied. The regular set of acceptability dimensions used and information collected in NuvaRing^®^ acceptability studies did not include measures of frequency of sexual encounters or sexual satisfaction. Sexual satisfaction investigated most of the time in separate studies using female sexuality indexes or other similar measures, showed mixed results. Sabatini & Cagliano pointed out that in their study the analysis of adverse events revealed that disturbance of sexual intercourse was a crucial point for acceptability, compliance and continuation (12).This is in line with other studies documenting the relationship between contraception and sexuality (59). Some authors believe that sexual side effects are the best predictors of discontinuation of oral contraceptives among heterosexual adult women (60). Actually contraceptives can affect women’s sexuality in a wide variety of ways beyond sexual functioning alone, for example, they can affect communication between sexual partners and women’s empowerment (61). Interestingly, as qualitative data showed in Rwanda, enhanced couples’ communication (for instance because of CVR use and potential increased lubrification), contributed to the acceptability of the NuvaRing^®^. The use of female sexual function indexes as well as aspects related to sexual relationship may help to improve our understanding of the relationships between contraception and sexuality, including for CVRs.

Overall, limited information was provided on actual experiences of women using CVR and cultural context which may affect CVR use (55). Further documenting actual women’s experiences using the CVR, and male partner opinions (including with respect to relationship and sexuality), can contribute to a better understanding of acceptability of and satisfaction with CVR (55). Awareness of the multiple contexts in ring users’ experience and giving a strong voice to women regarding their perception of contraceptive methods may inform the development and promotion approaches for CVR and more broadly vaginal rings (29,62,63).

This review has several limitations. First, given the lack of standardized definitions of acceptability and satisfaction we may have missed articles documenting CVR acceptability or satisfaction that were not *explicitly* using this terminology and instead referred to continuation or adherence, which was not a specific outcome of interest in this review. Second, we could not always deduct from the methods sections of included papers whether interviews included open-ended questions. This may have led to under-recording of the use of semi-structured interviews. However in-depth qualitative techniques, such as in-depth interviews or focus group discussions, were always clearly described in papers.

## Conclusion - recommendation

Many studies using mostly quantitative structured questionnaires have documented acceptability and satisfaction of hormonal CVRs, particularly NuvaRing^®^. Despite the use of similar dimensions in a number of studies, there was a lack of standardized definitions of acceptability and satisfaction. Sexual satisfaction or pleasure was not typically included in acceptability dimensions and findings were not very informative in terms of actual experiences of women using CVRs and the cultural context that may affect sexuality and contribute to shape acceptability of CVRs. The use of mixed-methods or qualitative approaches, including information on experiences of women using CVRs, relationship aspects, male partner opinions, and contextual sexual norms, may lead to a better understanding of acceptability and satisfaction of CVRs. In addition, the use of theoretical acceptability frameworks highlighting the actual pathway from acceptability to satisfaction and adherence might also be useful.

## Data Availability

All data relevant for the study are included in the paper or uploaded as supplementary information.

## Declaration of interests

The authors declare no conflicts of interest.

## Funding

This systematic review did not include primary data collection and no specific grant or funding was required.

## Contributorship statement

Conceived and designed the study: TD, VJ, LB, JvdW. Conducted data based searches: TD, VJ, LB. Conducted data screening and selected the studies: TD, VJ. Conducted data extraction: TD. Performed data analysis and presentation of results: TD, VJ. Wrote manuscript: TD, VJ, LB, JvdW. All authors provided input in manuscript writing and approved the final manuscript.

## Ethical statement

This review did not include primary data collection, and there was no patient and public involvement in the development of the protocol for the review.

## Data sharing

All data relevant for the study are included in the paper or uploaded as supplementary information.

## Notes

### Competing Interest Statement

The authors have declared no competing interest.

## References

1. Brache V, Faundes A. Contraceptive vaginal rings: a review. Contraception. 2010;82(5):418–27.

2. Monteiro I, Guazzelli C BL. Advances in contraceptive vaginal rings: what does the future hold? Expert Opin Pharmacother. 2018;

3. Baeten JM, Palanee-Phillips T, Brown ER, Schwartz K, Soto-Torres LE, Govender V, et al. Use of a Vaginal Ring Containing Dapivirine for HIV-1 Prevention in Women. NEnglJMed. 375(22):2121–32.

4. Trussell J. Contraceptive failure in the United States. Contraception. 2011;83(5):397–404.

5. Novak A, de la Loge C, Abetz L. Development and validation of an acceptability and satisfaction questionnaire for a contraceptive vaginal ring, NuvaRing((R)). Pharmacoeconomics. 2004;22(4):245–56.

6. Colwell HH, Mathias SD, Cimms TA, Rothman M, Friedman AJ, Patrick DL. The ORTHO BC-SAT--a satisfaction questionnaire for women using hormonal contraceptives. QualLife Res. 2006 Dec;15(10):1621–31.

7. Van Der Straten A, Montgomery ET, Cheng H, Wegner L, Masenga G, Von Mollendorf C, et al. High acceptability of a vaginal ring intended as a microbicide delivery method for HIV prevention in African women. AIDS Behav. 2012 Oct;16(7):1775–86.

8. Ahrendt H-JJ, Nisand I, Bastianelli C, Gomez MA, Gemzell-Danielsson K, Urdl W, et al. Efficacy, acceptability and tolerability of the combined contraceptive ring, NuvaRing, compared with an oral contraceptive containing 30 micrograms of ethinyl estradiol and 3 micrograms of drospirenone. Contraception. 2006 Dec;74(6):451–7.

9. Battaglia C, Morotti E, Persico N, Battaglia B, Busacchi P, Casadio P, et al. Clitoral vascularization and sexual behavior in young patients treated with drospirenone-ethinyl estradiol or contraceptive vaginal ring: a prospective, randomized, pilot study. J Sex Med. 2014 Feb;11(2):471–80.

10. Miller L, Verhoeven CHJ, Hout J. Extended regimens of the contraceptive vaginal ring: a randomized trial. ObstetGynecol. 2005 Sep;106(0029-7844 (Print)):473–82.

11. Mishell DR, Lumkin M, Stone S, Mishell Jr. DR. Inhibition of ovulation with cyclic use of progestogen-impregnated intravaginal devices. Am J Obstet Gynecol. 1972;113(7):927–32.

12. Sabatini R, Cagiano R. Comparison profiles of cycle control, side effects and sexual satisfaction of three hormonal contraceptives. Contraception. 2006 Sep;74(3):220–3.

13. Schafer JE, Osborne LM, Davis AR, Westhoff C. Acceptability and satisfaction using Quick Start with the contraceptive vaginal ring versus an oral contraceptive. Contraception. 2006;73(5):488–92.

14. Sivin I, Mishell DR, Alvarez F, Brache V, Elomaa K, Lahteenmaki P, et al. Contraceptive vaginal rings releasing Nestorone and ethinylestradiol: a 1-year dose-finding trial. Contraception. 2005 Feb;71(2):122–9.

15. Thiery M, Vandekerckhove D, Dhont M, Vermeulen A, Decoster JM. The medroxyprogesterone acetate intravaginal silastic ring as a contraceptive device. Contraception. 1976;13(5):605–17.

16. Weisberg E, Fraser IS, Mishell DRJ, Lacarra M, Bardin CW. The acceptability of a combined oestrogen/progestogen contraceptive vaginal ring. Contraception. 1995 Jan;51(1):39–44.

17. Stewart FH, Brown BA, Raine TR, Weitz TA, Harper CC. Adolescent and young women’s experience with the vaginal ring and oral contraceptive pills. J Pediatr Adolesc Gynecol [Internet]. 2007;20(6):345–51. Available from: http://www.ncbi.nlm.nih.gov/pmc/articles/PMC3163239/pdf/nihms227867.pdf

18. Gill K, Happel AU, Pidwell T, Mendelsohn A, Duyver M, Johnson L, et al. An open-label, randomized crossover study to evaluate the acceptability and preference for contraceptive options in female adolescents, 15 to 19 years of age in Cape Town, as a proxy for HIV prevention methods (UChoose). J Int AIDS Soc. 2020;23(10):1–10.

19. Caruso S, Cianci S, Panella M, Giunta G, Matarazzo MG, Cianci A. Comparative randomized study on the sexual function and quality of life of women on contraceptive vaginal ring containing ethinylestradiol/etonogestrel 3.47/11.00mg or 2.7/11.7mg. Gynecol Endocrinol [Internet]. 2019;0(0):1–5. Available from: https://doi.org/10.1080/09513590.2019.1603290

20. Creinin MD, Meyn LA, Borgatta L, Barnhart K, Jensen J, Burke AE, et al. Multicenter comparison of the contraceptive ring and patch: A randomized controlled trial. Obstet Gynecol. 2008;111(2):267–77.

21. Elaut E, Buysse A, De SP, De CG, Gerris J, Deschepper E, et al. Relation of androgen receptor sensitivity and mood to sexual desire in hormonal contraception users. Contraception. 2012 May;85(5):470–9.

22. Gilliam ML, Neustadt A, Kozloski M, Mistretta S, Tilmon S, Godfrey E. Adherence and acceptability of the contraceptive ring compared with the pill among students: A randomized controlled trial. Obstet Gynecol. 2010;115(3):503–10.

23. Gracia CR, Sammel MD, Charlesworth S, Lin H, Barnhart KT, Creinin MD. Sexual function in first-time contraceptive ring and contraceptive patch users. FertilSteril. 2010 Jan;93(1):21–8.

24. Guida M, Sardo AD, Bramante S, Sparice S, Acunzo G, Tommaselli GA, et al. Effects of two types of hormonal contraception - oral versus intravaginal - on the sexual life of women and their partners. Hum Reprod. 2005 Apr;20(4):1100–6.

25. Kestelyn E, Van Nuil JI, Umulisa MM, Umutoni G, Uwingabire A, Mwambarangwe L, et al. High acceptability of a contraceptive vaginal ring among women in Kigali, Rwanda. PLoSOne. 2018;13(6):e0199096..

26. Merkatz RB, Plagianos M, Hoskin E, Cooney M, Hewett PC, Mensch BS. Acceptability of the nestorone(R)/ethinyl estradiol contraceptive vaginal ring: development of a model; implications for introduction. Contraception. 2014 Nov;90(5):514–21.

27. Brucker C, Karck U, Merkle E. Cycle control, tolerability, efficacy and acceptability of the vaginal contraceptive ring, NuvaRing®: Results of clinical experience in Germany. Eur J Contracept Reprod Heal Care. 2008;13(1):31–8.

28. Buckshee K, Kumar S, Saraya L. Contraceptive vaginal ring--a rising star on the contraceptive horizon. Adv Contracept. 1990 Sep;6(3):177–83.

29. McLellan-Lemal E, Ondeng’e K, Gust DA, Desai M, Otieno FO, Madiega PA, et al. Contraceptive vaginal ring experiences among women and men in Kisumu, Kenya: A qualitative study. Front Womens Heal. 2017;2(1).

30. Merki-Feld GS. & Hund M. Clinical experience with NuvaRing® in daily practice in Switzerland: Cycle control and acceptability among women of all reproductive ages. Eur J Contracept Reprod Heal Care. 2007;12(3):240–7.

31. Merki-Feld GS, Hund M. Clinical experience with the combined contraceptive vaginal ring in Switzerland, including a subgroup analysis of previous hormonal contraceptive use. Eur J Contracept Reprod Heal Care. 2010 Dec;15(6).

32. Novák A, de la Loge C, Abetz L, van der Meulen EA, Novak A, de la Loge C, et al. The combined contraceptive vaginal ring, NuvaRing®: An international study of user acceptability. Contraception. 2003 Mar;67(3):187–94.

33. Pandit SN, Chauhan AR, Anagani M, Reddy S, Birla A, Ray SK. Multicenter Study of Contraceptive Vaginal Ring (NuvaRing®) in Normal Daily Practice in Indian Women. J Obstet Gynecol India. 2014 Dec;64(6):409–16.

34. Peipert JF, Zhao Q, Allsworth JE, Petrosky E, Madden T, Eisenberg D, et al. Continuation and Satisfaction of Reversible Contraception. Obstet Gynecol. 2011;117(5):1105–13.

35. RamaRao S, Clark H, Rajamani D, Ishaku S, Mané B, Obaré F, et al. Progesterone Vaginal Ring: Results of a Three-Country Acceptability Study. 2015.

36. Roumen FJ, Apter D, Mulders TMT, Dieben TOM. Efficacy, tolerability and acceptability of a novel contraceptive vaginal ring releasing etonogestrel and ethinyl oestradiol. HumReprod. 2001;16(3):469–75.

37. Roumen FJME, Op Ten Berg MMT, Hoomans EHM. The combined contraceptive vaginal ring (NuvaRing®): First experience in daily clinical practice in The Netherlands. Eur J Contracept Reprod Heal Care. 2006;11(1):14–22.

38. Sahota J, Barnes PMFF, Mansfield E, Bradley JL, Kirkman RJEE. Initial UK experience of the levonorgestrel-releasing contraceptive intravaginal ring. Adv Contracept. 1999;15(4):313–24.

39. Caruso S, Cianci S, Malandrino C, Cicero C, Lo Presti L, Cianci A, et al. Quality of sexual life of women using the contraceptive vaginal ring in extended cycles: Preliminary report. EurJContraceptReprodHealth Care. 2014 Aug;19(4):307–14.

40. Sanchez S, Araya, Araya C, Tijero M, Diaz S. Women’s perceptions and experience with the progesterone vaginal ring for contraception during breastfeeding. In London, England, Reproductive Health Matters, 1997.; p. 49–57. Available from: http://www.who.int/reproductive-health/publications/beyond_acceptability_users_perspectives_on_contraception/sanchez.en.pdf

41. Schindler AE. The 3-keto-desogestrel / ethinylestradiol ring: a new parenteral form of hormonal contraception. Eur J Obstet Gynecol Reprod Biol. 1993;49(1–2):p13–4.

42. Soni A, Garg S, Bangar R. Efficacy, user acceptability, tolerability, and cycle control of a combined contraceptive vaginal ring: The indian perspective. J Obstet Gynecol India. 2013 Oct;63(5):337–41.

43. Spencer BE, Jones V, Elstein M. The acceptability of the contraceptive vaginal ring. Br J Fam Plann. 1986;12(3):82–7.

44. Dam P, Paul J, Banerjee I, Sorkhel A, Chakravorty PS. To Study the Acceptability and Efficacy of Combined Contraceptive Vaginal Ring Amidst Indian Women. J Evol Med Dent Sci [Internet]. 2015;4(38):6602–12. Available from: http://www.jemds.com/data_pdf/3_PurvitaDam…….meena…….swe.pdf

45. Elder MG, Lawson JP, Elstein M, Nuttall ID. The efficacy and acceptability of a low-dose levonorgestrel intravaginal ring for contraception in a UK cohort. Contraception. 1991 Feb;43(2):129–37.

46. Faundes A, Hardy E, Reyes Q, Pastene L, Portes-Carrasco R. Acceptability of the contraceptive vaginal ring by rural and urban population in two Latin American Countries. Contraception. 1981;24(4):393–414.

47. Fine PM, Tryggestad J, Meyers NJ, Sangi-Haghpeykar H. Safety and acceptability with the use of a contraceptive vaginal ring after surgical or medical abortion. Contraception. 2007;75(5):367–71.

48. Guida M, Cibarelli F, Troisi J, Gallo A, Palumbo AR, Sardo ADS. Sexual life impact evaluation of different hormonal contraceptives on the basis of their methods of administration. ArchGynecolObstet. 2014 Dec;290(6):1239–47.

49. Hardy EE, Reyes Q, Gomez F, Portes-Carrasco R, Faundes A. User’s perception of the contraceptive vaginal ring: a field study in Brazil and the Dominican Republic. Stud Fam Plann. 1983 Nov;14(11):284–90.

50. Koetsawang S, Gao J, Krishna U, et al, World Health Organisation. Microdose intravaginal levonorgestrel contraception: a multicentre clinical trial. II. Expulsions and removals. Contraception. 1990;41(2):125–41.

51. Epstein LB, Sokal-Gutierrez K, Ivey SL, Raine T, Auerswald C. Adolescent Experiences with the Vaginal Ring. J Adolesc Heal. 2008 Jul;43(1):64–70.

52. Guida M, Di Carlo C, Troisi J, Gallo A, Cibarelli F, Martini E, et al. The sexuological impact of hormonal contraceptives based on their route of administration. Gynecol Endocrinol [Internet]. 2016 Dec 2 [cited 2017 Jan 31];1–5. Available from: http://www.ncbi.nlm.nih.gov/pubmed/27908210

53. Lete I, Doval JL, Perez-Campos E, Lertxundi R, Correa M, de V la, et al. Self-described impact of noncompliance among users of a combined hormonal contraceptive method. Contraception. 2008 Apr;77(4):276–82.

54. Novak A, de la Loge C, Abetz L. Development and validation of an acceptability and satisfaction questionnaire for a contraceptive vaginal ring, NuvaRing((R)). Pharmacoeconomics [Internet]. 2004;22(4):245–56. Available from: http://web.a.ebscohost.com.central.ezproxy.cuny.edu:2048/ehost/detail/detail?vid=2&sid=b01fdcbc-bebb-48f4-b3bd-230f186c7482%40sessionmgr4007&hid=4104&bdata=JnNpdGU9ZWhvc3QtbGl2ZQ%3D%3D#AN=12324639&db=a9h

55. Vargas SE, Midoun MM, Guillen M, Getz ML, Underhill K, Kuo C, et al. A Qualitative Systematic Review of Women’s Experiences Using Contraceptive Vaginal Rings: Implications for New Technologies. Perspect Sex Reprod Health [Internet]. 2019 Jun 20 [cited 2019 Nov 20];51(2):71–80. Available from: https://onlinelibrary.wiley.com/doi/abs/10.1363/psrh.12103

56. Griffin JB, Ridgeway K, Montgomery E, Torjesen K, Clark R, Peterson J, et al. Vaginal ring acceptability and related preferences among women in low- and middle-income countries: A systematic review and narrative synthesis. PLoS One. 2019;14(11):e0224898.

57. Roumen FJME, Mishell DRJ. The contraceptive vaginal ring, NuvaRing ®, a decade after its introduction. Eur J Contracept Reprod Heal Care [Internet]. 2012 Dec 31 [cited 2019 Jan 14];17(6):415–27. Available from: http://www.ncbi.nlm.nih.gov/pubmed/23113828

58. Morrow KM, Rosen RK, Salomon L, Woodsong C, Severy L, Fava JL, et al. Using integrated mixed methods to develop behavioral measures of factors associated with microbicide acceptability [Internet]. Vol. 21, Qual.Health Res. p. 987–99. Available from: http://pdf

59. Trussell J, Westoff CF. Contraceptive practice and trends in coital frequency. Fam Plann Perspect [Internet]. 1980 [cited 2019 Jan 14];12(5):246–9. Available from: http://www.ncbi.nlm.nih.gov/pubmed/7439349

60. Sanders SA, Graham CA, Bass JL, Bancroft J. A prospective study of the effects of oral contraceptives on sexuality and well-being and their relationship to discontinuation. Vol. 64, Contraception. 2001. p. 51–8.

61. Higgins JA, Smith NK. The Sexual Acceptability of Contraception: Reviewing the Literature and Building a New Concept. J Sex Res [Internet]. 2016;53(4–5):417–56. Available from: http://dx.doi.org/10.1080/00224499.2015.1134425

62. Hardon A. The development of contraceptive technologies: a feminist critique. Gend Dev [Internet]. 1994 Jun 20 [cited 2019 Jan 25];2(2):40–4. Available from: http://www.ncbi.nlm.nih.gov/pubmed/12345531

63. Woodsong C, Musara P, Chandipwisa A, Montgomery E, Alleman P, Chirenje M, et al. Interest in multipurpose prevention of HIV and pregnancy: perspectives of women, men, health professionals and community stakeholders in two vaginal gel studies in southern Africa. BJOG. 2014;121 Suppl:45–52.

