## Supplementary Material S1 for "Acceptability and satisfaction of contraceptive vaginal rings in clinical studies: a systematic review and narrative synthesis"

**Supplementary Material S1: Search strategy - Till December 31th 2020**

### Databases search

1. **Pubmed**

**Main Syntax Search : Contraceptive vaginal ring AND acceptability OR satisfaction OR sexual satisfaction):**
("contraceptive ring"[All Fields]) OR ("NuvaRing"[All Fields]) OR ("nuvaring"[All Fields]) OR ("contraceptive vaginal ring"[All Fields]) OR ("intravaginal ring"[All Fields]) OR ("progesterone"[MeSH Terms] AND ring) OR ("progesterone"[All Fields] AND ring) OR (("breast feeding"[MeSH Terms]) OR ("breast feeding"[All Fields]) OR ("breastfeeding"[All Fields]) AND (ring) OR (Contraceptive AND ring)) AND (acceptability[All Fields] OR ("personal satisfaction"[MeSH Terms]) OR ("personal"[All Fields] AND "satisfaction"[All Fields]) OR "orgasm"[MeSH Terms] OR ("sexual satisfaction"[All Fields]) OR (behavior AND sexual) OR ("orgasm"[All Fields]) OR (sexuality[MeSH Terms]) OR ("sexual"[All Fields] AND life[MeSH Terms]) OR (sexual AND "life"[All Fields]))

**Additional Syntax Search (Secondary objective - Methods)**
**Contraception, acceptability, satisfaction and Trials:**
**Search** ("contraception"[MeSH Terms] OR "contraceptive agents"[MeSH Terms] OR "contraception"[All Fields] OR "birth control"[All Fields] OR "family planning"[All Fields]) AND (acceptability[All Fields] OR "patient satisfaction"[MeSH Terms] OR "personal satisfaction"[MeSH Terms]) AND (trial[MeSH Terms])

Search (("contraception"[MeSH Terms] OR "contraceptive agents"[MeSH Terms]) OR "contraception"[All Fields]) OR "birth control"[All Fields])) AND (acceptability[All Fields] OR "patient satisfaction"[MeSH Terms] OR "personal satisfaction") AND ((qualitative) OR ("mixed-methods"))

2. **Web of knowledge:**

contraception * “vaginal ring” * acceptability
+ contraception * “vaginal ring” * satisfaction
+ contraception + vaginal ring * “sexual satisfaction”
+ contraception * “intravaginal ring” * acceptability
+ contraception * “intravaginal ring” * satisfaction

3. **CINAHL**

(contraceptive ring OR NuvaRing OR nuvaring OR intravaginal ring OR progesterone ring OR nesterone ring) AND (acceptability OR satisfaction OR orgasm OR sexuality OR sexual life)
